# Delayed Coal Plant Retirements in the Era of Data Centers Carry a Measurable Mortality Cost

**DOI:** 10.64898/2026.09.29.26364336

**Authors:** Suhwan Bong, Gianluca Guidi, Michael Cork, Lucas Henneman, Francesca Dominici

## Abstract

Coal-fired power plants remain a major source of fine particulate matter (PM_2.5_) exposure and mortality in the United States. Announced retirement dates mark when those burdens are expected to end, but rising electricity demand, partly from data centers supporting cloud computing and artificial intelligence, has led utilities and regulators to revisit them. Of 50 coal plants scheduled in January 2024 to retire before 2035, 20 had later or removed dates by January 2026. Projecting coal PM_2.5_ and mortality among Medicare beneficiaries from the 20 plants for 2026–2035, we estimate the revised schedule would result in 1,256 additional deaths (1,136 to 1,376), raising mortality to 3.1 times the original burden. Eighteen plants operate in balancing authorities with hyperscale data-center demand, and federal orders and utility filings supporting several plants’ operation cited data-center growth. Retirement schedules are revised faster than health consequences are assessed, and those delays carry a measurable mortality cost.

## Introduction

Coal-fired power plants have long been a major source of fine particulate matter (PM_2.5_) air pollution in the United States (*1–3*). Coal PM_2.5_ carries a mortality risk 2.1 times that of PM_2.5_ from all sources combined, and from 1999 to 2020 it contributed to an estimated 460,000 deaths among Medicare beneficiaries (*1*). Coal combustion emits primary particles and gaseous precursors, including sulfur dioxide and nitrogen oxides, that contribute to secondary PM_2.5_ formation and can affect populations far from the generating facility (*4–6*). These exposures are associated with increased risk of mortality and other adverse health outcomes (*7–11*). The resulting health impacts vary across regions and population groups because of the geography of power plants, atmospheric transport, and residential settlement patterns (*1, 12*). Although U.S. coal generation has declined substantially (*13*), 208 coal-fired power plants were still listed as operating in the United States in the January 2026 EIA-860M inventory.

The decline of coal-fired electricity generation has been among the major drivers of air quality improvements in the United States over the past several decades (*14, 15*). Plant retirements, reduced coal dispatch, and installation of emissions controls have contributed to large reductions in sulfur dioxide, nitrogen oxides, primary particulate matter, and coal PM_2.5_ (*16, 17*). These changes have generated corresponding health benefits for exposed populations (*14, 18*). Announced retirement schedules therefore encode expectations about when future emissions, exposure, and health burdens will cease. When a retirement is postponed, the anticipated air quality and health gains are postponed as well.

Electricity demand in the United States is projected to increase after a prolonged period of relatively slow growth, with data centers supporting cloud computing and artificial intelligence among the major sources of new large loads (*19–21*). U.S. data centers consumed an estimated 176 TWh of electricity in 2023, and their annual electricity use was projected to reach 325 to 580 TWh by 2028 (*20*). The North American Electric Reliability Corporation has identified data centers and other large commercial and industrial loads as major drivers of projected demand growth and emerging challenges for resource-adequacy planning (*21*). Rapid connection of these loads can increase near-term capacity and reliability requirements, particularly where the expansion of generation and transmission is constrained by long development and interconnection timelines (*21–23*). Energy Information Administration scenarios further indicate that data-center demand could increase near-term fossil-fuel generation (*24*).

At the same time, federal regulatory requirements for fossil fuel–fired power plants have been reopened or revised, including greenhouse gas standards for fossil fuel–fired electric generating units and Mercury and Air Toxics Standards for coal- and oil-fired electric utility steam generating units (*25, 26*). Together, these broader electricity-system and regulatory changes may lead utilities, regulators, and grid operators to revisit previously announced retirement dates, making their public health consequences a timely question.

Existing coal-health research has estimated mortality risks and historical health burdens attributable to coal PM_2.5_ (*1*), characterized inequities in exposure to coal-fired power plant emissions (*12*), and evaluated the health benefits of past coal emissions reductions, plant retirements, retrofits, or fuel conversions (*14, 27, 28*). Little is known about the potential negative health consequences if the retirement of some coal-fired power plants is delayed.

Because coal PM_2.5_ can be transported over long distances, delayed retirements can increase exposure not only near the plant but also in downwind and far-away communities. Thus, the populations bearing the health burden of continued coal operation may be geographically distinct from the plant-host community and from the regions receiving reliability or electricity-supply benefits (*12, 29, 30*). Quantifying the spatial distribution of additional exposure to coal PM_2.5_ and its mortality effects is therefore necessary to evaluate not only the magnitude but also the equity implications of delayed retirements. Because a relatively small number of facilities can account for a large share of power-sector air pollution damages, even a limited number of retirement delays may produce substantial negative impacts on health (*31–34*).

In this paper, we quantify the additional mortality associated with the documented delays in coal plant retirement dates in the United States. We compared coal plant retirement plans reported in the January 2024 U.S. Energy Information Administration Form EIA-860M generator inventory (original plans) with revised plans reported in the January 2026 inventory (*35*). Among 50 coal plants with reported retirement dates before 2035 in the January 2024 inventory, 20 had retirement dates that were delayed or removed by January 2026. For these 20 plants, we estimated annual plant-specific coal PM_2.5_ exposure at the ZIP Code Tabulation Area (ZCTA) level under the original and revised retirement scenarios for 2026–2035. Throughout, coal PM_2.5_ refers to the SO_2_-derived secondary component and excludes primary particulate matter and nitrate formed from NO_x_, so the estimates are conservative with respect to total coal-attributable PM_2.5_. We then estimated the projected increase in mortality among Medicare beneficiaries by comparing the two scenarios. We also evaluated alternative emissions and operating duration assumptions in sensitivity analyses in Supplementary materials.

This analysis provides a generator-level framework for translating revisions to retirement schedules into additional coal PM_2.5_ exposure and attributable mortality. We harmonize generator-level retirement dates, unit-level SO_2_ emissions, and outputs of a reduced-complexity atmospheric model to estimate spatially resolved exposure and mortality across 31,473 ZCTAs, to identify which facilities drive the burden, and to examine connections between the affected plants and hyperscale data-center demand.

## Results

Historically, coal PM_2.5_ exposure was concentrated in the southern and eastern United States, particularly in major coal-generating and downwind regions, including the Ohio Valley, Appalachia, the Midwest, and parts of the Southeast (fig. S1). This spatial pattern reflects both the location of coal-fired power plants and the long-range atmospheric transport of coal PM_2.5_. Population-weighted historical exposure also varied across demographic groups, with the largest departure from the overall population mean observed among Black populations (fig. S2).

### Revisions to announced coal retirement plans

The retirement schedules of the coal-fired power plants changed substantially between the January 2024 and January 2026 EIA-860M inventories. Among 50 coal plants with reported retirement dates before 2035 in the January 2024 inventory, 20 plants, or 40%, had retirement dates that were either delayed or removed by January 2026 (fig. 1 and table S1). Of these 20 plants, 10 had delayed retirement dates and 10 no longer had a reported retirement date. Because EIA-860M reports retirement dates per generator, a plant was classified as affected if at least one coal-fired generator had a later or removed date; 34 of the 44 coal generators at these plants were affected, and the affected share of plant SO_2_ ranged from 5% at Sherburne County (MN) to 100% at 12 plants (table S1). These revised-retirement plants were distributed across multiple regions of the United States and included facilities with both near-term and later scheduled retirement dates.

**Fig. 1.**
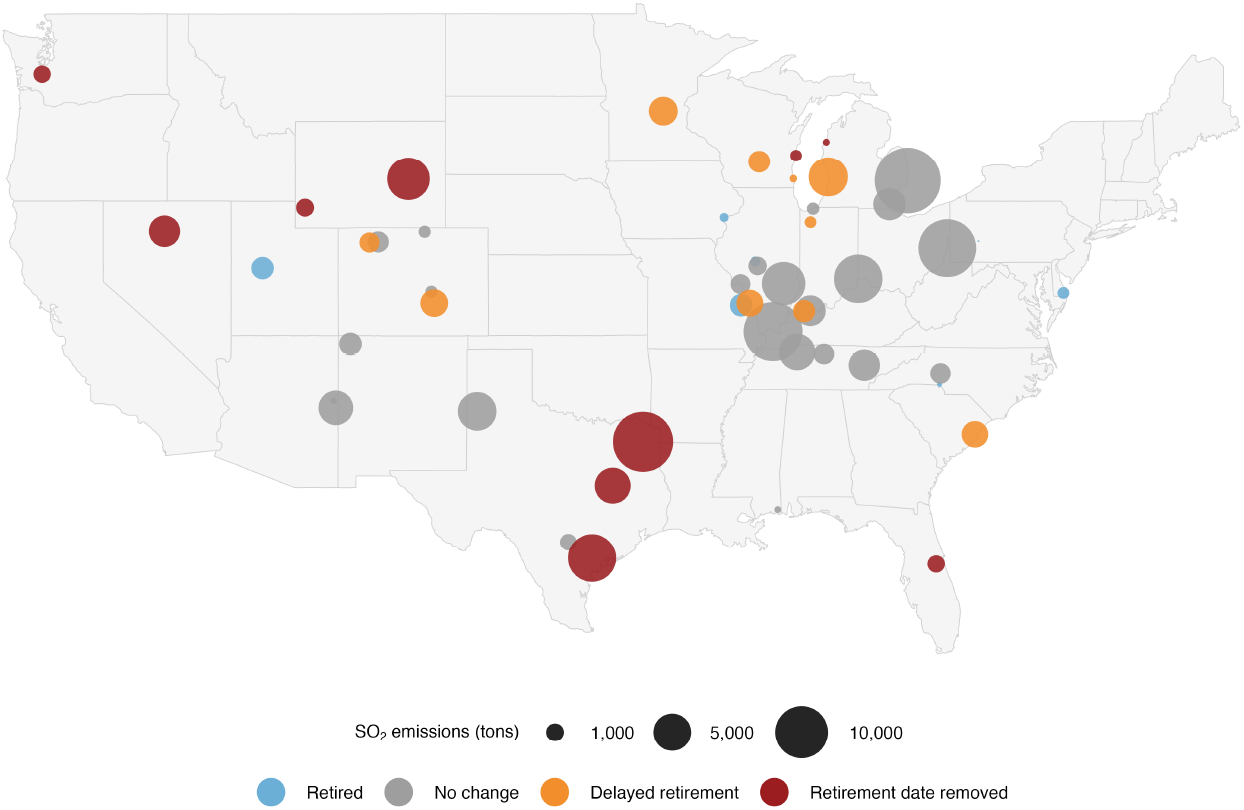
Revisions to announced coal plant retirement schedules between January 2024 and January 2026. Map of 50 U.S. coal plants with reported retirement dates before 2035 in the January 2024 EIA-860M inventory. Point colors indicate retirement status in the January 2026 EIA-860M inventory: retired, no change, delayed retirement, or retirement date removed. Point size is proportional to 2024 SO_2_ emissions. The 20 plants classified as delayed retirement or retirement date removed define the revised retirement plant set used in the main health impact analysis.

The revised retirement plant set included plants originally scheduled to retire as early as 2025 as well as plants scheduled for retirement later in the decade. Some plants retained a reported retirement date but shifted that date later, whereas others no longer had a reported retirement date in the January 2026 inventory. The share of eligible plants with later or removed retirement dates was higher in 2024–2026 than in the preceding comparison periods (table S2). These changes indicate that announced retirement dates are not fixed planning endpoints; instead, they may be revised over short periods in ways that alter expectations about future coal operation and emissions.

Under the revised retirement schedule, the affected plants would remain available for coal-fired generation longer than under the original schedule. For some plants, the revision represented a discrete extension of one or several years. For others, removal of the retirement date created a larger divergence from the original schedule. Because health damages from coal-fired power generation arise from continued emissions rather than from retirement announcements themselves, even short delays can preserve exposure that would otherwise have been avoided.

### Delayed retirements preserve coal PM_2.5_ exposure

Delayed retirement dates preserved substantial coal PM_2.5_ exposure over 2026–2035. Under the original retirement scenario, coal PM_2.5_ from the 20 affected plants declined as plants retired according to the January 2024 schedule. Under the revised retirement scenario, these exposure reductions were delayed or removed, producing cumulative additional coal PM_2.5_ exposure (fig. 2). The magnitude and spatial distribution of this additional exposure reflected plant location, recent SO_2_ emissions, plant-specific coal PM_2.5_ transport patterns, and the duration of continued operation. When expressed as a share of cumulative coal PM_2.5_ exposure from the 20 plants under the revised scenario, the additional exposure attributable to retirement-schedule changes accounted for a substantial proportion of projected coal PM_2.5_-years across many ZCTAs (fig. S3). The revised scenario also increased cumulative population-weighted coal PM_2.5_ exposure across all groups examined (fig. S4). Increases ranged from 187% among Asian populations to 244% among American Indian and Alaska Native populations, whose historical exposure had been the lowest of any group (fig. S2).

**Fig. 2.**
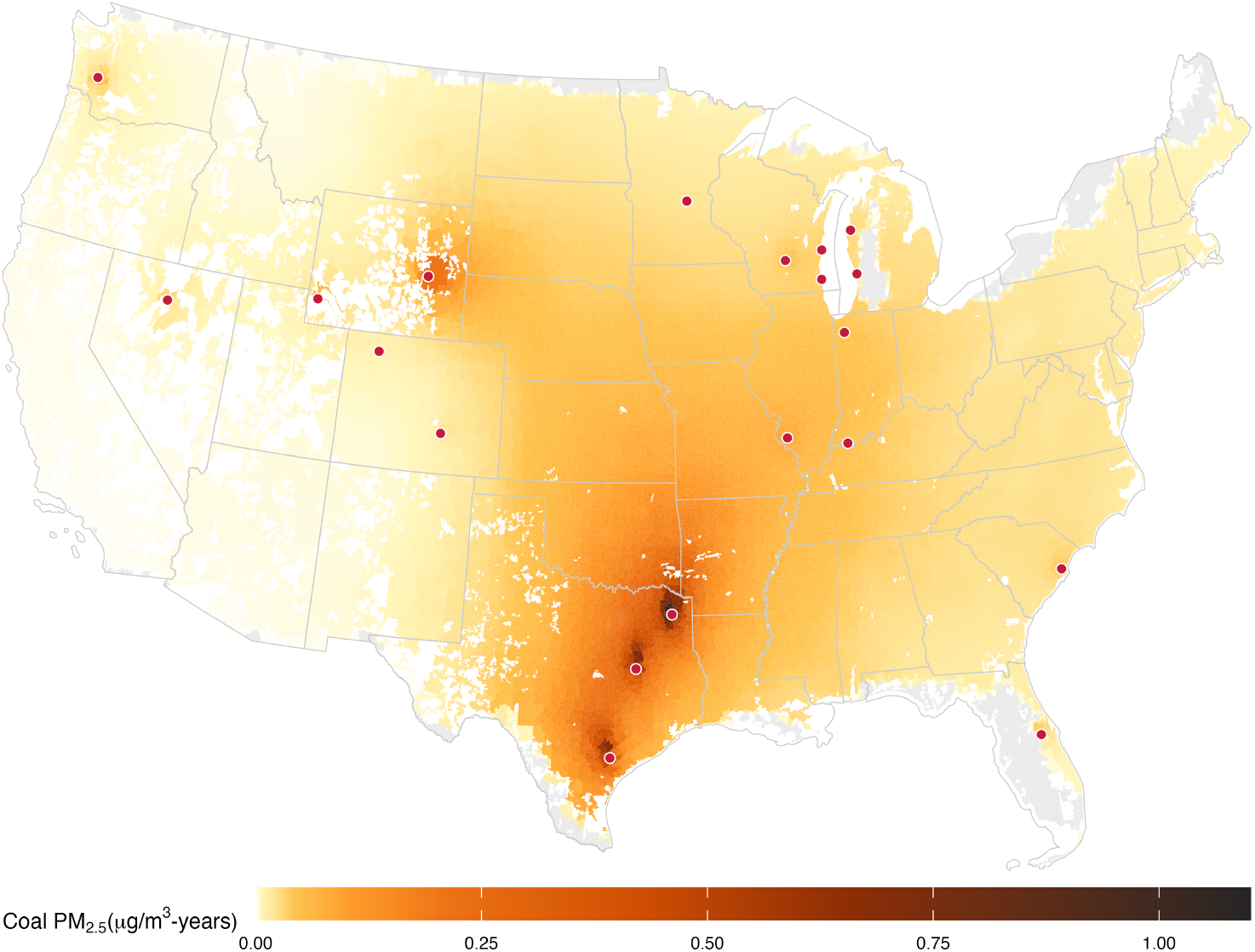
Cumulative additional coal PM_2.5_ exposure from delayed coal plant retirements compared to the original scenario, 2026–2035. ZCTA-level cumulative additional coal PM_2.5_ exposure under the revised retirement scenario relative to the original retirement scenario. Colors show the cumulative difference in coal PM_2.5_ exposure between the revised and original scenarios over 2026–2035. Points indicate coal plants with delayed retirement dates. Values are reported in micrograms per cubic meter–years. Gray areas indicate ZCTAs without an estimated historical plant-ZCTA fixed effect for the affected plants; white areas fall outside Census ZCTA coverage.

The largest cumulative exposure increases were concentrated in the southern Plains, especially in Texas and Oklahoma and surrounding areas, where several plants with delayed or removed retirement dates had large projected exposure impacts. Plant-specific exposure maps for all 20 affected plants showed substantial heterogeneity in the magnitude and geographic extent of additional coal PM_2.5_ exposure (fig. S5). Additional coal PM_2.5_ was not confined to communities immediately surrounding the affected plants. Instead, exposure extended across broad downwind areas, consistent with long-range transport of secondary particulate matter from coal-fired power plant emissions. This spatial pattern indicates that the health consequences of retirement delays depend not only on which plants remain online, but also on where their emissions are transported and which populations lie within their exposure footprints.

### Delayed retirements substantially increase attributable mortality

Figure 3 shows the total number of excess deaths in the Medicare population when comparing cumulative exposure to coal PM_2.5_ between the revised and original retirement scenarios. The two mortality trajectories diverged over the projection period. Under the original retirement scenario (blue line), the total number of deaths increased early but then grew more slowly as plants retired according to the January 2024 schedule. Under the revised retirement scenario, the total number of deaths continues to increase because plants with delayed retirement dates were assumed to remain available for coal-fired operation for additional years.

**Fig. 3.**
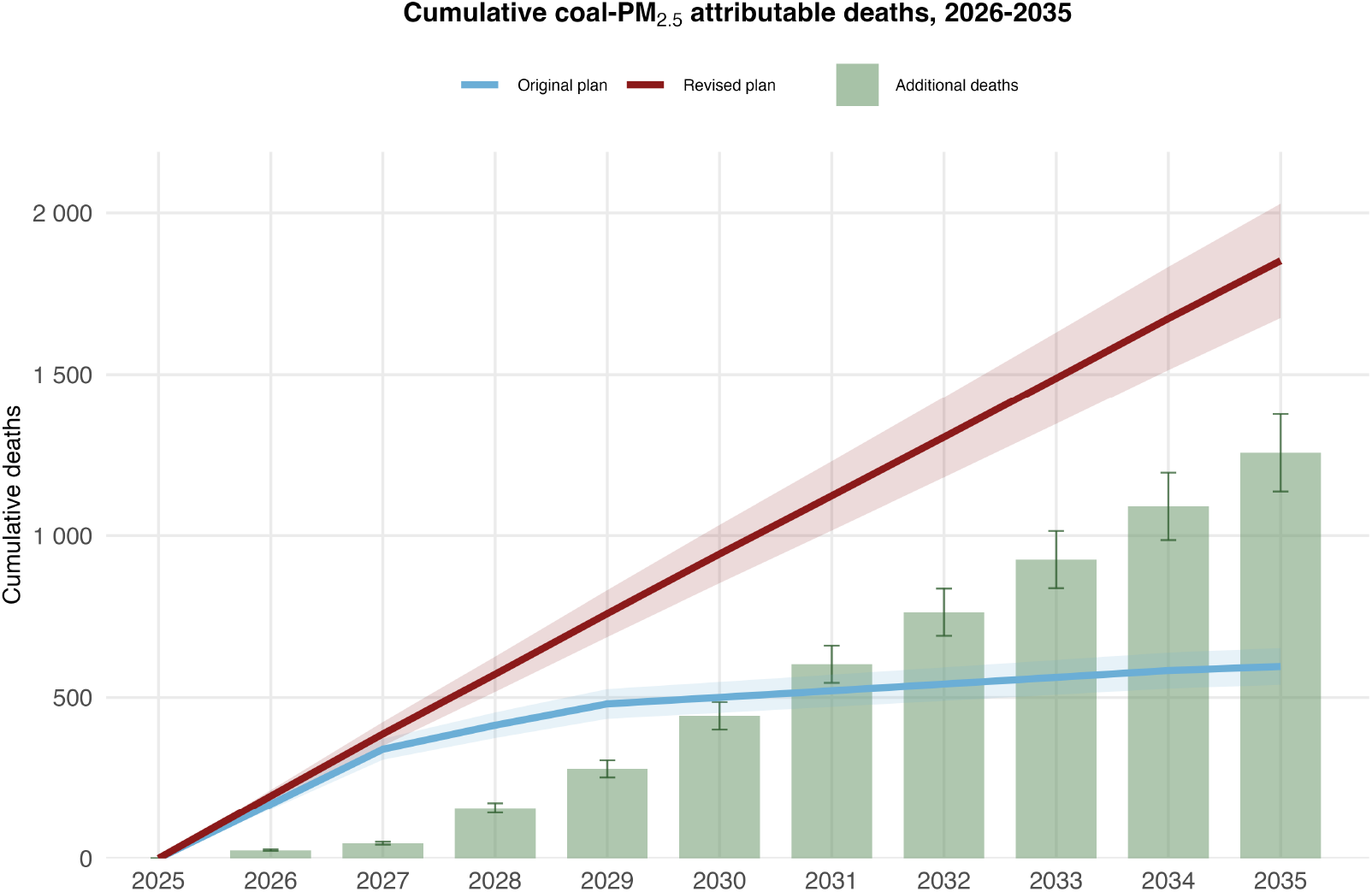
Cumulative all-cause deaths among Medicare beneficiaries attributable to coal PM_2.5_ from the 20 plants with delayed retirement dates. Blue and red lines show cumulative attributable deaths under the original and revised retirement scenarios, respectively. Shaded bands show estimates based on lower and upper coal PM_2.5_ mortality-response estimates. Green bars show cumulative additional deaths under the revised scenario relative to the original scenario; error bars show corresponding lower and upper estimates that reflect uncertainty in the coal PM_2.5_ concentration-response function.

By 2035, the revised retirement scenario was projected to result in 1,256 (1,136 to 1,376) additional deaths among Medicare beneficiaries compared to the original scenario. Cumulative attributable mortality was 1,852 deaths under the revised scenario and 596 under the original scenario, a ratio of 3.11.

The temporal pattern of mortality highlights the importance of retirement timing. Delayed retirements do not produce a one-time increase in exposure; each additional year of continued operation adds another year of coal-related PM_2.5_ exposure and attributable mortality. As a result, the cumulative health burden grows over time. These results indicate that the health gains expected from announced coal retirements are contingent on whether those retirements occur as planned.

### Plant-level attribution identifies large source-specific mortality burdens

Plant-level attribution showed that the projected national mortality burden was concentrated among identifiable source-level retirement changes. For plants with later or removed retirement dates, the revised retirement schedules produced substantial additional mortality among exposed Medicare beneficiaries over 2026–2035 (fig. 4 and table S3). Welsh (TX) was projected to contribute 435 additional Medicare deaths, followed by Coleto Creek (TX) with 316 additional deaths and Limestone (TX) with 279 additional deaths. Dave Johnston (WY) contributed 99 additional deaths. These four largest contributors together accounted for 90% of the total.

**Fig. 4.**
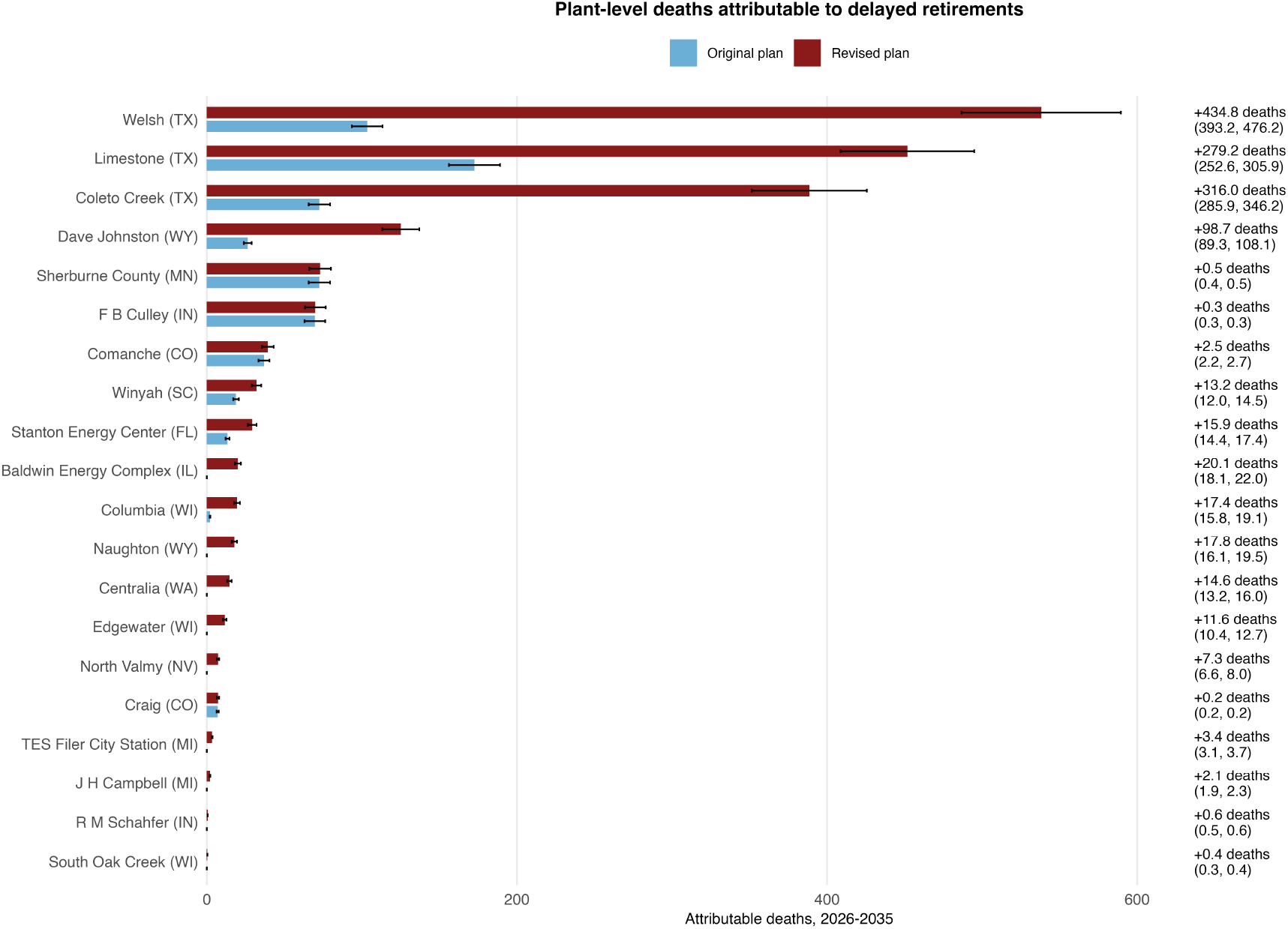
Plant-level mortality burdens attributable to delayed retirement, 2026–2035. Cumulative all-cause deaths among Medicare beneficiaries attributable to coal PM_2.5_ from coal plants with delayed retirement, summed over 2026–2035. Blue bars show deaths under the original retirement scenario and red bars show deaths under the revised retirement scenario.

Error bars show lower and upper estimates based on the confidence limits of the coal PM_2.5_ concentration-response function. Text labels show additional deaths under the revised scenario relative to the original scenario.

Together, these estimates show that delayed retirement dates generated source-specific public health burdens. The plant-level analysis complements the national cumulative estimate by translating aggregate excess mortality into burdens attributable to individual facilities, with Welsh (TX), Coleto Creek (TX), and Limestone (TX) accounting for approximately 1,030 of the 1,256 additional deaths, or about 82% of the total. These three largest contributors also produced broad plant-specific exposure footprints across Texas and neighboring downwind areas (fig. 5). Plant-specific mortality estimates varied with the duration of continued operation, plant emissions, and historical atmospheric transport toward populated areas, producing regional exposure patterns that extended beyond communities near the plants.

**Fig. 5.**
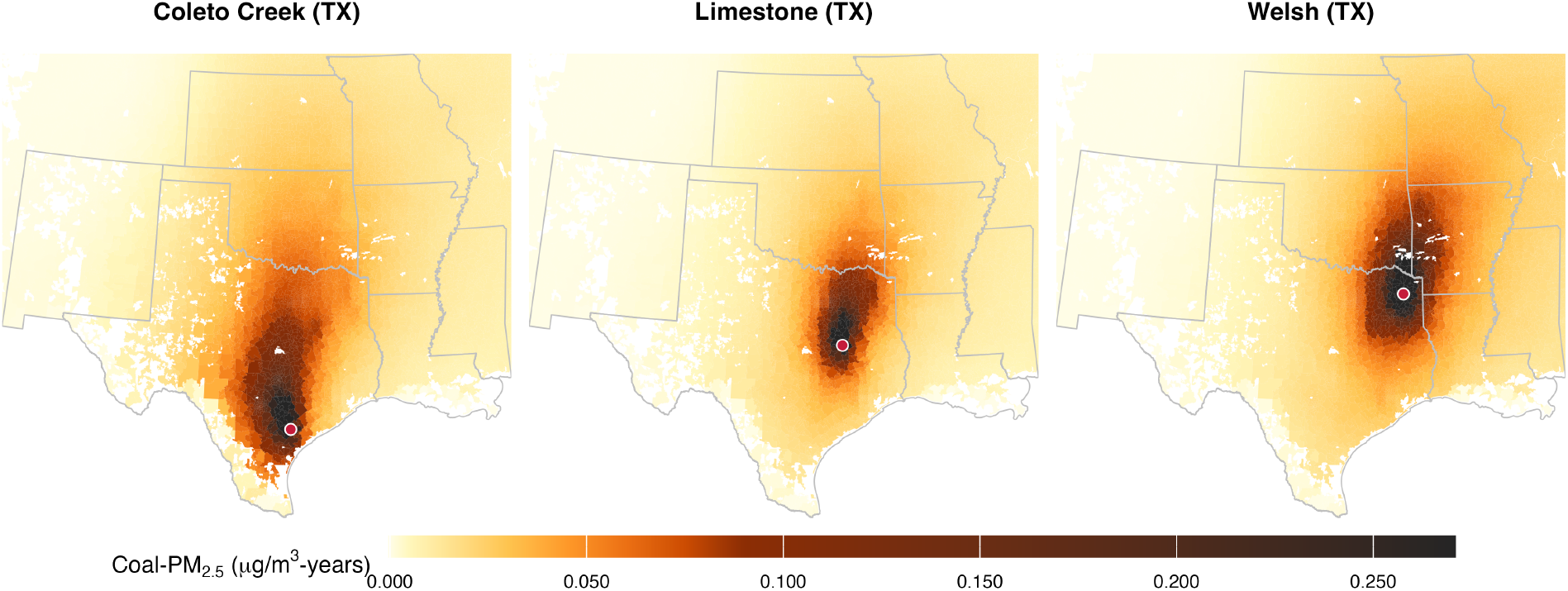
Additional coal PM_2.5_ exposure attributable to delayed retirement for Coleto Creek (TX), Limestone (TX), and Welsh (TX), 2026–2035. Maps show cumulative additional coal PM2.5 exposure over 2026–2035 under the revised retirement scenario relative to the original retirement scenario. Values are summed across projection years at the ZCTA level and reported in micrograms per cubic meter–years. These three plants account for 82% of the projected additional mortality.

### Hyperscale data-center demand and the affected plants

Using the attributional framework of (*36*), which estimates hyperscale data-center (HDC) electricity demand within each balancing authority and attributes that demand to power plants in proportion to their positive annual net generation, we matched the 20 plants with delayed or removed retirement dates to EIA plant identifiers and EPA eGRID2023 Revision 2 balancing-authority assignments (*37*). Eighteen of the 20 plants were located in seven balancing authorities containing positive HDC electricity demand (table S8). This classification indicates regional overlap between HDC electricity demand and the generation mix that included the affected plants.

Official records and contemporaneous reporting linked the continued operation of several affected plants to growing electricity demand from data centers. During the comparison period, Department of Energy emergency orders directed Campbell (MI), Centralia (WA), Craig (CO), Culley (IN), and Schahfer (IN) to remain available on resource-adequacy grounds and cited the rapid expansion of AI and data centers among the demand pressures increasing the need to retain dispatchable generation (*38–44*). The Campbell continuation orders provided the most specific connection, citing Consumers Energy’s plan to supply 1 GW to a planned hyperscale data center and its reported momentum with data centers within a 9-GW development pipeline (*39, 40*).

Additional evidence showed similar demand pressures at other plants: the co-owners of Columbia (WI) cited customer growth and a grid-operator recommendation to monitor large-load additions and delay resource retirements; the operator of South Oak Creek (WI) cited tightened regional energy supply and high electricity demand; and Xcel Energy sold land near the Sherburne County (MN) plant to Microsoft for a planned data center (*45–47*). Federal orders directing Stanton (FL) to remain available cited projected data-center and other large-load growth in Florida, including eight planned data centers in the state (*48, 49*). Together, these records indicate that data-center demand contributed to the planning and reliability conditions supporting the continued operation of several affected plants.

## Discussion

This study shows that revisions to coal plant retirement schedules lead to additional exposure to coal PM_2.5_ and substantial excess mortality. Among coal plants with announced retirement dates before 2035, 40% had retirement dates that were delayed or removed between the January 2024 and January 2026 EIA-860M inventories. Under the revised retirement scenario, cumulative coal PM2.5 attributable mortality among Medicare beneficiaries was projected to be 1,256 additional deaths over 2026–2035, 3.11 times the original scenario burden. The magnitude of the estimated burden is notable because our analysis captures only a documented subset of an ongoing and evolving retirement planning process. Additional retirement delays have been publicly discussed or announced but are not yet reflected in EIA-860M reporting, and other delays may occur after January 2026 or affect plants outside this comparison. Moreover, our estimates capture only coal PM_2.5_ attributable mortality among Medicare beneficiaries and do not include other pollutants, younger populations, or nonfatal health outcomes. Even within this constrained and documented plant set, the projected health burden was substantial.

It is concerning that 18 of the 20 plants with delayed or removed retirement dates operate in balancing authorities with hyperscale data-center demand, and that the continuation orders for one plant pointed to a utility’s plan to supply 1 GW to a single planned hyperscale facility (*39, 40*). Because hyperscale campuses can add gigawatt-scale load within a few years, faster than replacement generation and transmission can be permitted, interconnected, and built (*21–23*), aging coal units risk becoming the default bridge resource for this demand. If that pattern holds, the mortality costs of powering hyperscale computing would fall largely on downwind communities that receive little of its economic benefit and have no voice in data-center siting or power-procurement decisions.

These findings also extend prior evidence on the health burden of coal-fired electricity generation by linking historical coal PM_2.5_ exposures to prospective retirement scenarios (*50*). Previous work has shown that coal PM_2.5_ imposed large historical mortality burdens in the United States and that those burdens declined as coal generation and emissions fell (*1, 12, 14, 18*). Building on this evidence, we used the historical relationship between plant-level SO_2_ emissions and plant-specific coal PM_2.5_ exposure to project future exposure and mortality under alternative retirement schedules.

Plant-level attribution showed that much of the national mortality burden arose from a small number of facilities, including Welsh (TX), Coleto Creek (TX), Limestone (TX), and Dave Johnston (WY). By identifying the small number of facilities responsible for most of the projected burden, this plant-level attribution makes the results more actionable than aggregate national burden estimates.

Several limitations should be considered. First, this is a scenario-based analysis of reported retirement plans, not a prediction of actual future dispatch. Plant operations may change with fuel prices, electricity demand, reliability events, regulatory decisions, maintenance schedules, and replacement-resource availability. Second, future emissions were represented using each generator’s mean annual SO_2_ emissions during 2023–2025 combined with scenario-specific calendar-availability fractions. This approach isolates the effect of retirement timing but does not model future changes in dispatch intensity, pollution-control performance, coal sulfur content, or non-SO_2_ emissions. Third, future coal PM_2.5_ estimates rely on historical plant-specific exposure fields and emissions-scaling assumptions. They therefore do not fully capture future year-to-year variation in meteorology, atmospheric chemistry, or emissions controls, which are recognized sources of uncertainty in future air quality projections (*51, 52*). Fourth, mortality estimates were limited to all-cause mortality among Medicare beneficiaries. We did not estimate mortality among younger populations or nonfatal outcomes such as cardiovascular events, respiratory morbidity, asthma exacerbations, hospitalizations, or adverse birth outcomes (*18, 53*). However, the concentration–response function used for mortality estimation was derived specifically for coal PM_2.5_ exposure among Medicare beneficiaries, matching both the exposure and population evaluated here and reducing potential mischaracterization of attributable mortality. ZCTAs without an estimated plant–ZCTA fixed effect are assigned no exposure. These contain 10% of Medicare beneficiaries for most plants, and 23% for Stanton (FL) and 20% for Winyah (SC), so the estimates are conservative in this respect. Finally, we did not model replacement generation or broader power-system responses. Our estimates therefore represent the health burden preserved by continued operation of the delayed retirement plants, not the full net health effect of all generation changes that might occur under the original retirement pathway (*30, 54*). Under the original schedule, retired capacity would have been partly replaced by gas generation or by increased dispatch at other coal plants, both of which would make the net effect smaller than the gross effect reported here. In addition, the data-center analysis identifies regional overlap and documentary context rather than causal attribution.

These results have implications for how reliability and public health are considered in electricity-system planning. Federal orders retaining several of these plants cited data-center growth as part of their reliability rationale. As data-center electricity demand continues to grow, such demand pressures may continue to shape decisions about when coal plants retire. Reliability concerns are real, especially in regions facing demand growth, resource adequacy constraints, transmission limitations, interconnection delays, and climate change-driven increases in extreme weather events. However, keeping coal plants online is not a health-neutral reliability strategy. Continued coal operation preserves emissions of PM_2.5_ precursors and associated mortality risks that can extend well beyond nearby communities. Because retirement schedules effectively determine when plant-specific emissions and associated health burdens end, revisions to those schedules should be evaluated not only in terms of reliability, replacement capacity, and system cost, but also for their public health consequences.

## Materials and Methods

### Study design and retirement scenarios

We obtained coal plant retirement schedules from the U.S. Energy Information Administration Preliminary Monthly Electric Generator Inventory based on Form EIA-860M (*35*). We used the January 2024 inventory to define the original retirement schedule and the January 2026 inventory to define the revised retirement schedule. We restricted the main analysis to coal plants with at least one operating coal-fired unit that reported a retirement date before 2035 in the January 2024 inventory.

We matched generators across inventories using EIA plant and generator identifiers. Because EIA-860M reports retirement dates per generator, we retained the generator as the unit of analysis rather than aggregating dates to the plant level: collapsing to a single plant date either erases a delay that applies to one generator or applies it to the whole plant’s emissions. We classified each coal-fired generator as retired, no change, delayed retirement, or retirement date removed, and classified a plant as affected when at least one of its coal-fired generators had a later or removed date. Among 50 eligible coal plants, 20 met this criterion; of the 44 coal-fired generators at these plants, 19 had delayed dates, 15 had dates removed, 7 were unchanged, 2 had already retired, and 1 was converted to natural gas (table S1).

For each generator, year, and retirement scenario, we constructed a calendar-availability fraction to prorate emissions in a partial retirement year. We assigned a value of 1 in full calendar years before the scenario-specific retirement date, 0 after retirement, and the number of months before retirement divided by 12 in the retirement year. Under the original retirement scenario, we used dates from January 2024. Under the revised retirement scenario, we used later dates from January 2026 and assigned a fraction of 1 through 2035 to generators with removed dates. By multiplying full-year average emissions 2023-2025 by this fraction, we avoided representing a within-year retirement as either a full year of emissions or no emissions.

Generator-level fractions were combined into a plant-level availability fraction weighted by each generator’s 2023–2025 mean SO2. We weighted by emissions rather than by capacity because scrubber status and utilization differ across generators within a plant.

### SO_2_ emissions

We obtained annual SO_2_ emissions from the U.S. Environmental Protection Agency Clean Air Markets Program Data (*57*): plant-level emissions for 1999–2025, used to fit the historical model, and unit-level emissions for 2023–2025 at the 20 affected plants, used to weight and prorate the projections. For each generator, we defined the full year emissions baseline for 2026-2035 as the mean of annual SO_2_ emissions during 2023-2025. For the 2026-2035 projections, we held this recent three-year average fixed until the scenario-specific retirement date and multiplied it by the calendar-availability fraction described above. Under the original retirement scenario, we set emissions to zero after the retirement date reported in the January 2024 EIA-860M inventory. Under the revised retirement scenario, we set emissions to zero after the later retirement date reported in the January 2026 inventory for plants with delayed retirement dates.

### Coal PM_2.5_ Exposure modeling

Coal PM_2.5_ exposure estimates were based on the long-term coal PM_2.5_ source-impact dataset. These data provide annual unit-specific coal PM_2.5_ concentrations associated with SO_2_ emissions from U.S. coal-fired power plants during 1999-2020 (*1, 56*). The HYSPLIT with Average Dispersion model (HyADS), a reduced-complexity atmospheric model, generated the source-impact fields. We aggregated the gridded fields to ZCTAs and linked them to annual plant-level SO_2_ emissions. Throughout the manuscript and Supplementary Materials, coal PM_2.5_ refers to this SO_2_-associated component and does not include all primary PM_2.5_ or PM_2.5_ formed from other precursors.

Because we did not have future HyADS source-impact fields for 2026-2035, we estimated an empirical relationship between historical plant-level SO_2_ emissions and ZCTA-level coal PM_2.5_ and applied that relationship to scenario-specific future emissions. This approach follows reduced-complexity source-receptor modeling, which links source-specific emissions to receptor-level PM_2.5_ concentrations for policy-relevant scenario analysis (*59, 60*). We used a log-log specification to represent the relationship between SO2 emissions and coal PM2.5 as multiplicative and to obtain positive concentration predictions after transformation. In this formulation, *γ* is an elasticity; for a given plant-ZCTA pair, a 1% change in SO2 emissions corresponds to an estimated *γ* % change in coal PM2.5. This application assumes that each plant-ZCTA source-receptor relationship estimated from 1999–2020 remains applicable during 2026– 2035.

To project coal PM_2.5_ under future retirement scenarios, we modeled the historical relationship between plant-level SO_2_ emissions and ZCTA-level coal PM_2.5_. For plant *p*, ZCTA *z*, and year *t*, we fit the log-log model

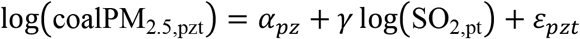

where coalPM_2.5,pzt_ is the coal PM_2.5_ concentration from plant *p* in ZCTA z and year *t*, SO_2,*pt*_ is annual SO_2_, emissions from plant *p* in year *t*, α_*pz*_ is a plant-ZCTA fixed effect capturing the average source-receptor relationship between each plant and ZCTA, gamma is the elasticity of coal PM_2.5_ with respect to SO_2_ emissions, and ε_*pzt*_ is the residual error term. This specification implies that, for a given plant-ZCTA pair in year *t* and *t*^∗^, the ratio of predicted coal PM_2.5_ concentrations under two emissions levels equals the ratio of SO_2_ emissions raised to gamma.

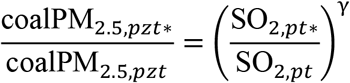

We excluded observations with zero coal PM_2.5_ concentration or zero SO_2_ emissions before log transformation. We fit the model by ordinary least squares with standard errors clustered by plant using all data from 1999 to 2020. The fitted model showed strong explanatory performance, with an adjusted R^2^ of 0.945 and a within R^2^ of 0.834. The estimated SO_2_ elasticity was 1.165 (95% CI, 1.124 to 1.206, table S4), indicating that coal PM_2.5_ increased slightly more than proportionally with SO_2_ emissions in the fitted historical data.

The HyADS source-impact fields are constructed by scaling dispersion weights by emissions and calibrating the result, so γ partly recovers that construction. Because α is estimated over 1999–2020, when emissions were higher than in 2023–2025 at every affected plant (ratios 0.03 to 0.68), γ > 1 makes the downward extrapolation steeper and the projection more conservative: refitting with γ fixed at 1 raises the estimate from 1,256 to 1,427 additional deaths and lowers the share attributable to the three Texas plants from 82% to 80%.

We used the fitted model to predict coal PM2.5 under the original and revised retirement scenarios. Exponentiating a fitted log-scale value yields the conditional median rather than the conditional mean, so predicted concentrations were multiplied by a Duan smearing factor, which requires no distributional assumption:

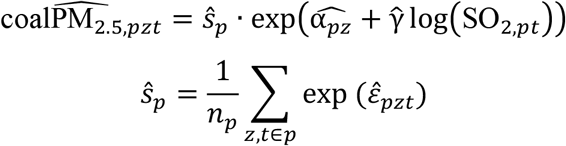

Where *n*_*p*_ is the number of observations for plant *p* and 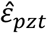 is a log-scale residual. Residual variance differs across plants (standard deviation 0.35 to 0.63), so the factor was computed separately for each plant rather than pooled; it ranges from 1.063 to 1.226, with a pooled value of 1.169 (table S4). The estimator drops singleton plant–ZCTA groups, leaving 12,474,346 observations across 616,801 pairs.

For each plant and year from 2026 to 2035, we used scenario-specific SO2 emissions to predict plant-ZCTA coal PM2.5 under each scenario. We calculated annual ZCTA-level coal PM2.5 exposure under each scenario by summing predicted contributions from the 20 plants with delayed or removed retirement dates. For each ZCTA and year, we calculated additional coal PM2.5 exposure attributable to delayed or removed retirement dates as the revised scenario minus the original scenario. Cumulative additional exposure over 2026-2035 was calculated by summing annual differences across the projection period.

Attributable deaths were computed from each ZCTA’s total coal PM2.5 across the 20 plants and then allocated to plants in proportion to their contribution to that total, rather than by summing plant-specific attributable fractions. The two approaches differ by 0.006% at these concentrations.

### Subgroup exposure analysis

We calculated population-weighted coal PM_2.5_ exposure by sex, race, and age group. We obtained ZCTA-level demographic population counts from 2020 U.S. Census Bureau demographic tabulations (*58*). These counts are used only to weight exposure by demographic subgroup and do not enter the mortality calculation, which uses Medicare enrollment and deaths throughout. For each subgroup, population-weighted exposure was calculated by weighting ZCTA-level coal PM_2.5_ concentrations by the subgroup population in each ZCTA.

For the historical analysis, we used long-term annual mean coal PM_2.5_ concentrations for 1999-2020 and calculated subgroup-specific population-weighted mean exposures. We calculated percent differences relative to the overall population-weighted mean. For the prospective retirement scenario analysis, we calculated cumulative population-weighted coal PM_2.5_ exposure under the original and revised retirement scenarios for 2026-2035. For each subgroup, we weighted cumulative exposure across ZCTAs by the subgroup. We calculated percent increases under the revised scenario as the revised-scenario cumulative population-weighted exposure minus the original scenario exposure, divided by the original-scenario exposure.

### Medicare population and baseline mortality rates

We obtained baseline ZCTA-level Medicare enrollment from Centers for Medicare & Medicaid Services data and national Medicare enrollment projections for 2026–2035 from the Medicare Trustees Report and accompanying supplementary tables (*61*). To construct projected ZCTA-level Medicare populations, we allocated national Medicare enrollment projections to ZCTAs according to each ZCTA’s share of Medicare enrollment in 2019. Thus, national enrollment changed by year, but we held its ZCTA distribution fixed at the 2019 pattern. National enrollment rises from 69.5 million in 2025 to 81.3 million in 2035, an increase of approximately 17% (table S5). Future geographic redistribution of Medicare beneficiaries could change local mortality estimates. We calculated baseline all-cause mortality rates from 2019 ZCTA-level Medicare deaths and enrollment and held the rates fixed across 2026–2035 (table S6). We used 2019 rather than 2020 because Medicare all-cause mortality in 2020 was elevated by the COVID-19 pandemic (0.04377, versus 0.03801 in 2019 and 0.03785 in 2023).

### Statistical analysis

We calculated all mortality estimates at the plant-ZCTA-year level and then aggregated across plants, ZCTAs, and years. For each plant *p*, ZCTA *z*, and year *t*, we estimated coal PM_2.5_ attributable deaths among Medicare beneficiaries using the model from (*1*). The calculation combined the ZCTA-level Medicare baseline mortality rate, projected Medicare enrollment, scenario-specific coal PM_2.5_ exposure, and the coal PM_2.5_ mortality-response coefficient from (*1*).

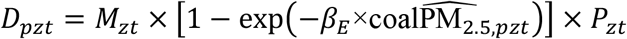

Where *D*_*pzt*_ is the number of all-cause deaths among Medicare beneficiaries attributable to coal PM_2.5_ from plant *p* in ZCTA *z*, and year *t*; *M*_*zt*_ is the baseline all-cause mortality rate among Medicare beneficiaries in ZCTA *z* and year *t*; *p*_*zt*_ is the projected number of Medicare enrollees in ZCTA *z* and year *t*; 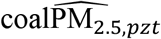 is the predicted coal PM_2.5_ concentration from plant *p* in ZCTA *z*, and year *t*; and β_E_ is the log-linear coal PM_2.5_ mortality-response coefficient. This formulation estimates the attributable fraction as 1 ™ exp(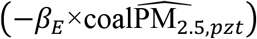), which is then multiplied by projected baseline deaths, *M*_*zt*_ *p*_*zt*_. For each scenario, we calculated cumulative coal PM_2.5_ attributable deaths over 2026-2035 by summing plant-specific attributable deaths across plants, ZCTAs, and years:

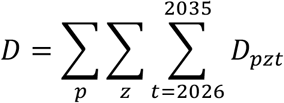

We calculated additional deaths attributable to delayed or removed retirement dates as the difference between the revised and original retirement scenarios. We calculated plant-specific additional deaths analogously for each plant. We also calculated the relative mortality burden under the revised retirement scenario as the ratio of cumulative attributable deaths under the revised scenario to cumulative attributable deaths under the original scenario. To calculate lower and upper mortality estimates, we used the lower and upper confidence limits for *β*_*E*_. The coal PM_2.5_ mortality-response coefficient corresponds to a relative risk of 1.0125 (1.0113 to 1.0137) per 1*μg/*M^3^, taken from (*1*).

These lower and upper estimates propagate only the confidence limits of *β*_*E*_. They do not incorporate uncertainty in the estimated SO2 elasticity γ, the plant–ZCTA fixed effects, the HyADS source-impact fields, future SO2 emissions, or baseline mortality rates. Uncertainty arising from future emissions and operating-duration assumptions is characterized separately through the scenario analyses in table S7.

### Sensitivity analyses of future SO_2_ emissions and retirement date assumptions

The main analysis required assumptions about both the annual SO_2_ emissions of plants that remained available to operate and the duration of operation for plants whose reported retirement dates had been removed. We used each generator’s mean annual SO_2_ emissions during 2023-2025 as the main active-year emissions baseline because these years represented the most recent emissions data available for the affected plants. Future emissions could differ from this recent mean if plants operate less or more intensively, undergo operational changes, or respond to changes in electricity demand. In addition, removal of a reported retirement date indicates that the previously announced date no longer applies but does not establish how long the plant will remain in operation.

We therefore conducted three complementary sensitivity analyses addressing distinct sources of uncertainty. The first evaluated proportional changes in future emissions across all affected plants. The second used generator-specific low and high emissions observed during the recent baseline period to account for differences in the recent operating range of individual plants. The third evaluated shorter durations of continued operation for plants whose retirement dates had been removed. Each analysis retained the historical plant-ZCTA fixed effects, estimated SO_2_-coal PM_2.5_ elasticity, Medicare population projections, ZCTA-specific baseline mortality rates, and coal PM_2.5_ mortality-response coefficients used in the main analysis. For the emissions sensitivity analyses, the alternative emissions baseline was applied to both the original and revised retirement scenarios.

### Uniform emissions-scaling scenarios

To evaluate the consequences of lower or higher future emissions, we repeated the analysis using 50%, 75%, 125%, and 150% of each generator’s mean annual SO_2_ emissions during 2023-2025. These scenarios characterize how the estimated burden changes under proportional deviations from the recent mean emissions baseline.

### Generator-specific emissions scenarios

Uniform scaling applies the same proportional change to every plant and does not represent differences in the recent operating range of individual facilities. We therefore conducted a separate analysis using the minimum and maximum annual SO_2_ emissions observed for each generator during 2023-2025, summed to the plant level. These values preserve plant-level heterogeneity and represent recently observed low- and high-emissions conditions. The generator-specific minimum and maximum emissions baselines were applied to both the original and revised retirement scenarios.

### Alternative assumptions for removed retirement dates

In the main analysis, plants whose reported retirement dates had been removed were assumed to remain available through the end of the 2026-2035 projection period. This assumption represents continued operation throughout the study period but may overstate operating duration if these plants retire after a shorter delay.

To evaluate the influence of this assumption, we assigned alternative revised retirement dates two years and five years after the original reported date, applied only to generators whose retirement date had been removed. Generators that retained a reported date kept it. Plants with explicitly reported revised retirement dates retained those dates, and the original retirement scenario retained the original reported dates for all plants. We applied the same calendar-availability fraction used in the main analysis, including partial-year availability during the assigned retirement year. These analyses retained the main plant-specific 2023-2025 mean SO_2_ emissions baseline.

## Data Availability

Code to support analyses in this work is documented on GitHub. CMS Medicare datasets, in accordance with the data use agreement, must be acquired from the Centers for Medicare & Medicaid Services. Coal PM2.5 source-impact fields are available from OSF. EIA-860M, EPA CAMPD, eGRID, and U.S. Census data are publicly available at the URLs cited in the manuscript.

## Data, code, and materials availability

Code to support analyses in this work is documented on GitHub (*55*). CMS Medicare datasets, in accordance with the data use agreement, must be acquired from the Centers for Medicare & Medicaid Services. Coal PM2.5 source-impact fields are available from (*56*). EIA-860M (*35*), EPA CAMPD (*57*), eGRID (*37*), and U.S. Census data (*58*) are publicly available at the URLs cited.

## References and Notes

1. L. Henneman, C. Choirat, I. Dedoussi, F. Dominici, J. Roberts, C. Zigler, Mortality risk from United States coal electricity generation. Science 382, 941–946 (2023).

2. G. K. Singh, P. Rajeev, D. Paul, T. Gupta, “Atmospheric Emissions from Thermal (Coal-Fired) Power Plants and Associated Environmental Impacts” in Pollutants from Energy Sources: Characterization and Control (Springer, 2018), pp. 53–72.

3. T. C. Odubo, E. A. Kosoe, “Sources of Air Pollutants: Impacts and Solutions” in Air Pollutants in the Context of One Health: Fundamentals, Sources, and Impacts (Springer, 2024), pp. 75–121.

4. C. Dedoussi, S. D. Eastham, E. Monier, S. R. Barrett, Premature mortality related to United States cross-state air pollution. Nature 578, 261–265 (2020).

5. L. R. Henneman, C. Choirat, C. Ivey, K. Cummiskey, C. M. Zigler, Characterizing population exposure to coal emissions sources in the United States using the HyADS model. Atmospheric Environment 203, 271–280 (2019).

6. N. Fann, K. R. Baker, C. M. Fulcher, Characterizing the PM2.5-related health benefits of emission reductions for 17 industrial, area and mobile emission sectors across the US. Environment International 49, 141–151 (2012).

7. X. Wu, D. Braun, J. Schwartz, M.-A. Kioumourtzoglou, F. Dominici, Evaluating the impact of long-term exposure to fine particulate matter on mortality among the elderly. Science Advances 6, eaba5692 (2020).

8. Q. Di, Y. Wang, A. Zanobetti, Y. Wang, P. Koutrakis, C. Choirat, F. Dominici, J. D. Schwartz, Air pollution and mortality in the Medicare population. New England Journal of Medicine 376, 2513–2522 (2017).

9. C. A. Pope III, R. T. Burnett, M. J. Thun, E. E. Calle, D. Krewski, K. Ito, G. D. Thurston, Lung cancer, cardiopulmonary mortality, and long-term exposure to fine particulate air pollution. JAMA 287, 1132–1141 (2002).

10. D. W. Dockery, C. A. Pope, X. Xu, J. D. Spengler, J. H. Ware, M. E. Fay, B. G. Ferris Jr., F. E. Speizer, An association between air pollution and mortality in six US cities. New England Journal of Medicine 329, 1753–1759 (1993).

11. World Health Organization, WHO Global Air Quality Guidelines: Particulate Matter (PM2.5 and PM10), Ozone, Nitrogen Dioxide, Sulfur Dioxide and Carbon Monoxide (World Health Organization, 2021).

12. L. R. Henneman, M. M. Rasel, C. Choirat, S. C. Anenberg, C. Zigler, Inequitable exposures to US coal power plant–related PM2.5: 22 years and counting. Environmental Health Perspectives 131, 037005 (2023).

13. U.S. Energy Information Administration, “U.S. Coal-Fired Electricity Generation Decreased in 2022 and 2023” (U.S. Energy Information Administration, 2024); https://www.eia.gov/todayinenergy/detail.php?id=62043.

14. L. R. F. Henneman, C. Choirat, C. M. Zigler, Accountability assessment of health improvements in the United States associated with reduced coal emissions between 2005 and 2012. Epidemiology 30, 477–485 (2019).

15. M. Filonchyk, M. P. Peterson, An integrated analysis of air pollution from US coal-fired power plants. Geoscience Frontiers 14, 101498 (2023).

16. U.S. Energy Information Administration, “Changes in Coal Sector Led to Less SO2 and NOx Emissions from Electric Power Industry” (U.S. Energy Information Administration, 2018); https://www.eia.gov/todayinenergy/detail.php?id=37752.

17. United States Environmental Protection Agency, “Progress Report—Emissions Reductions” (United States Environmental Protection Agency, 2024); https://www.epa.gov/power-sector/progress-report-emissions-reductions.

18. J. A. Casey, J. G. Su, L. R. Henneman, C. Zigler, A. M. Neophytou, R. Catalano, R. Gondalia, Y.-T. Chen, L. Kaye, S. S. Moyer, Improved asthma outcomes observed in the vicinity of coal power plant retirement, retrofit and conversion to natural gas. Nature Energy 5, 398–408 (2020).

19. U.S. Energy Information Administration, “EIA Forecasts Strongest Four-Year Growth in U.S. Electricity Demand Since 2000, Fueled by Data Centers” (U.S. Energy Information Administration, 2026); https://www.eia.gov/pressroom/releases/press582.php.

20. A. Shehabi, S. J. Smith, A. Hubbard, A. Newkirk, N. Lei, M. A. B. Siddik, B. Holecek, J. Koomey, E. Masanet, D. Sartor, “2024 United States Data Center Energy Usage Report” (LBNL-2001637, Lawrence Berkeley National Laboratory, 2024); 10.71468/P1WC7Q.

21. North American Electric Reliability Corporation, “2025 Long-Term Reliability Assessment” (North American Electric Reliability Corporation, 2026); https://www.nerc.com/globalassets/our-work/assessments/nerc_ltra_2025.pdf.

22. U.S. Department of Energy, “National Transmission Needs Study” (U.S. Department of Energy, 2023); https://www.energy.gov/oe/national-transmission-needs-study.

23. J. Rand, N. Manderlink, S. Zhang, C. Talley, W. Gorman, R. H. Wiser, J. Seel, J. M. Kemp, S. Jeong, F. Kahrl, “Queued Up: 2025 Edition, Characteristics of Power Plants Seeking Transmission Interconnection As of the End of 2024” (Lawrence Berkeley National Laboratory, 2025); https://emp.lbl.gov/queues.

24. U.S. Energy Information Administration, “Fossil Generation Could Rise with Faster-Than-Expected Growth in Data Center Power Demand” (U.S. Energy Information Administration, 2026); https://www.eia.gov/todayinenergy/detail.php?id=67344.

25. United States Environmental Protection Agency, “Greenhouse Gas Standards and Guidelines for Fossil Fuel-Fired Power Plants” (United States Environmental Protection Agency, 2025); https://www.epa.gov/stationary-sources-air-pollution/greenhouse-gas-standards-and-guidelines-fossil-fuel-fired-power.

26. United States Environmental Protection Agency, “Mercury and Air Toxics Standards” (United States Environmental Protection Agency, 2026); https://www.epa.gov/stationary-sources-air-pollution/mercury-and-air-toxics-standards.

27. J. A. Casey, D. Karasek, E. L. Ogburn, D. E. Goin, K. Dang, P. A. Braveman, R. Morello-Frosch, Retirements of coal and oil power plants in California: association with reduced preterm birth among populations nearby. American Journal of Epidemiology 187, 1586–1594 (2018).

28. M. Fan, Y. Wang, The impact of PM2.5 on mortality in older adults: evidence from retirement of coal-fired power plants in the United States. Environmental Health 19, 28 (2020).

29. A. L. Goodkind, C. W. Tessum, J. S. Coggins, J. D. Hill, J. D. Marshall, Fine-scale damage estimates of particulate matter air pollution reveal opportunities for location-specific mitigation of emissions. Proceedings of the National Academy of Sciences 116, 8775–8780 (2019).

30. C. Campos Morales, E. L. Pakhtigian, J. R. Landry, H. Wiseman, A. T. Pham, W. Peng, Designing retirement strategies for coal-fired power plants to mitigate air pollution and health impacts. Environmental Science & Technology 58, 15371–15380 (2024).

31. D. Tong, Q. Zhang, S. J. Davis, F. Liu, B. Zheng, G. Geng, T. Xue, M. Li, C. Hong, Z. Lu, Targeted emission reductions from global super-polluting power plant units. Nature Sustainability 1, 59–68 (2018).

32. N. Z. Muller, R. Mendelsohn, W. Nordhaus, Environmental accounting for pollution in the United States economy. American Economic Review 101, 1649–1675 (2011).

33. N. Maamoun, R. Kennedy, X. Jin, J. Urpelainen, Identifying coal-fired power plants for early retirement. Renewable and Sustainable Energy Reviews 126, 109833 (2020).

34. S. Gathrid, J. Wayland, S. Wayland, R. Deshmukh, G. C. Wu, Strategies to accelerate US coal power phase-out using contextual retirement vulnerabilities. Nature Energy 10, 1274–1288 (2025).

35. U.S. Energy Information Administration, Preliminary Monthly Electric Generator Inventory (based on Form EIA-860M as a supplement to Form EIA-860), U.S. Energy Information Administration (2026); https://www.eia.gov/electricity/data/eia860m.

36. G. Guidi, F. Dominici, T. Squartini, C. Sprinkle, J. Gilmour, K. Butler, E. Bell, S. Delaney, F. J. Bargagli-Stoffi, Assessing the carbon emissions and energy consumption of U.S. hyperscale data centers. arXiv:2606.05420 [cs.AI] (2026).

37. United States Environmental Protection Agency, eGRID2023 Revision 2, U.S. Environmental Protection Agency (2025); https://www.epa.gov/egrid/detailed-data.

38. U.S. Department of Energy, “Order No. 202-25-3” (U.S. Department of Energy, 2025).

39. U.S. Department of Energy, “Order No. 202-25-7” (U.S. Department of Energy, 2025).

40. U.S. Department of Energy, “Order No. 202-25-9” (U.S. Department of Energy, 2025).

41. U.S. Department of Energy, “Order No. 202-25-11” (U.S. Department of Energy, 2025).

42. U.S. Department of Energy, “Order No. 202-25-12” (U.S. Department of Energy, 2025).

43. U.S. Department of Energy, “Order No. 202-25-13” (U.S. Department of Energy, 2025).

44. U.S. Department of Energy, “Order No. 202-25-14” (U.S. Department of Energy, 2025).

45. Alliant Energy, “Wisconsin Energy Companies Announce Plan to Continue Operations at Columbia Energy Center, Commit to Exploring Conversion to Natural Gas” (Alliant Energy, 2024); https://www.alliantenergy.com/news/news-center/2024/12/120424-columbia-energy-center.

46. K. Marohn, “Xcel Energy Sells Land in Becker to Microsoft for Data Center” (MPR News, 2024); https://www.mprnews.org/story/2024/02/21/xcel-energy-sells-land-in-becker-to-microsoft-for-data-center.

47. We Energies, “We Energies Announces Updated Timeline for Oak Creek Plant Retirements” (We Energies, 2025); https://news.we-energies.com/we-energies-announces-updated-timeline-for-oak-creek-plant-retirements/.

48. U.S. Department of Energy, “Order No. 202-26-26” (U.S. Department of Energy, 2026).

49. U.S. Department of Energy, “Order No. 202-26-42” (U.S. Department of Energy, 2026).

50. M. M. Rasel, D. S. Cohan, D. Tong, L. R. Henneman, Emissions reductions at coal power plants continue to offer routes to meet new US PM2.5 standards. Environmental Science & Technology 60, 13796–13803 (2026).

51. D. J. Jacob, D. A. Winner, Effect of climate change on air quality. Atmospheric Environment 43, 51–63 (2009).

52. R. Doherty, F. O’Connor, S. Turnock, Projections of future air quality are uncertain. But which source of uncertainty is most important? Journal of Geophysical Research: Atmospheres 127, e2022JD037948 (2022).

53. R. D. Brook, S. Rajagopalan, C. A. Pope, J. R. Brook, A. Bhatnagar, A. V. Diez-Roux, F. Holguin, Y. Hong, R. V. Luepker, M. A. Mittleman, A. Peters, D. Siscovick, S. C. Smith, L. Whitsel, J. D. Kaufman, Particulate matter air pollution and cardiovascular disease. Circulation 121, 2331–2378 (2010).

54. T. M. Thompson, S. Rausch, R. K. Saari, N. E. Selin, A systems approach to evaluating the air quality co-benefits of US carbon policies. Nature Climate Change 4, 917–923 (2014).

55. S. Bong, G. Guidi, M. Cork, L. Henneman, F. Dominici, Code for: Delayed coal plant retirements in the era of data centers carry a measurable mortality cost, GitHub (2026); https://github.com/NSAPH-Projects/coal_retirement_delays-mortality.

56. L. R. F. Henneman, Long-term coal PM2.5 source impacts, OSF (2023); https://osf.io/8gdau/.

57. United States Environmental Protection Agency, Clean Air Markets Program Data, U.S. Environmental Protection Agency (2026); https://campd.epa.gov/.

58. U.S. Census Bureau, 2020 Census Demographic and Housing Characteristics File (DHC), U.S. Census Bureau (2023); https://www.census.gov/data/tables/2023/dec/2020-census-dhc.html.

59. C. W. Tessum, J. D. Hill, J. D. Marshall, InMAP: A model for air pollution interventions. PLOS ONE 12, e0176131 (2017).

60. United States Environmental Protection Agency, “Reduced-Form Tools for Calculating PM2.5 and Ozone Benefits” (United States Environmental Protection Agency, 2026); https://www.epa.gov/benmap/reduced-form-tools-calculating-pm25-and-ozone-benefits.

61. Boards of Trustees of the Federal Hospital Insurance and Federal Supplementary Medical Insurance Trust Funds, “The 2025 Annual Report of the Boards of Trustees of the Federal Hospital Insurance and Federal Supplementary Medical Insurance Trust Funds” (H. Doc. 119-63, U.S. Government Publishing Office, 2025); https://www.cms.gov/data-research/statistics-trends-and-reports/trustees-report-trust-funds.

