## Supplementary Materials for "Delayed Coal Plant Retirements in the Era of Data Centers Carry a Measurable Mortality Cost"

### Supplementary Text

Historical comparison of retirement-schedule revisions

We applied the same generator-level matching procedure to five consecutive two-year January EIA-860M inventory windows: 2016–2018, 2018–2020, 2020–2022, 2022–2024, and 2024–2026. For each baseline inventory, eligible plants were those with at least one operating coal-fired generator reporting a retirement year earlier than the baseline year plus 11. Eligible generators were matched to the January operating inventory two years later using plant and generator identifiers. The proportion of eligible plants with a later or removed retirement date was 21.2% in 2016–2018, 12.1% in 2018–2020, 20.8% in 2020–2022, 17.1% in 2022–2024, and 40.0% in 2024–2026 (table S2). Thus, the 2024–2026 window had the largest proportion among the five periods examined.

Sensitivity to future $SO_2$ emissions and retirement-date assumptions

Estimated cumulative additional Medicare mortality varied with the assumed level of future $SO_2$ emissions. Under the uniform emissions-scaling scenarios, setting emissions to 50% and 75% of each generator's 2023-2025 mean produced 560 additional deaths (lower-upper estimate, 507 to 614) and 898 additional deaths (813 to 984), respectively. These estimates were 55% and 28% lower than the main estimate of 1,256 additional deaths (1,136 to 1,376). Setting emissions to 125% and 150% of the recent mean produced 1,630 additional deaths (1,474 to 1,785) and 2,016 additional deaths (1,823 to 2,208), respectively, corresponding to increases of 30% and 60% relative to the main estimate. Using each generator's minimum annual SO2 emissions observed during 2023-2025 produced 922 additional deaths (834 to 1,009), 27% fewer than in the main analysis. Using each generator's maximum annual emissions produced 1,747 additional deaths (1,580 to 1,914), 39% more than in the main analysis. The assumed duration of continued operation was particularly influential for plants whose retirement dates had been removed. Assigning these plants retirement dates two years after their original dates produced 365 additional deaths (331 to 400), a reduction of 71% from the main estimate. Assigning retirement dates five years after the original dates produced 846 additional deaths (765 to 926), a reduction of 33% (fig. S6 and table S7).

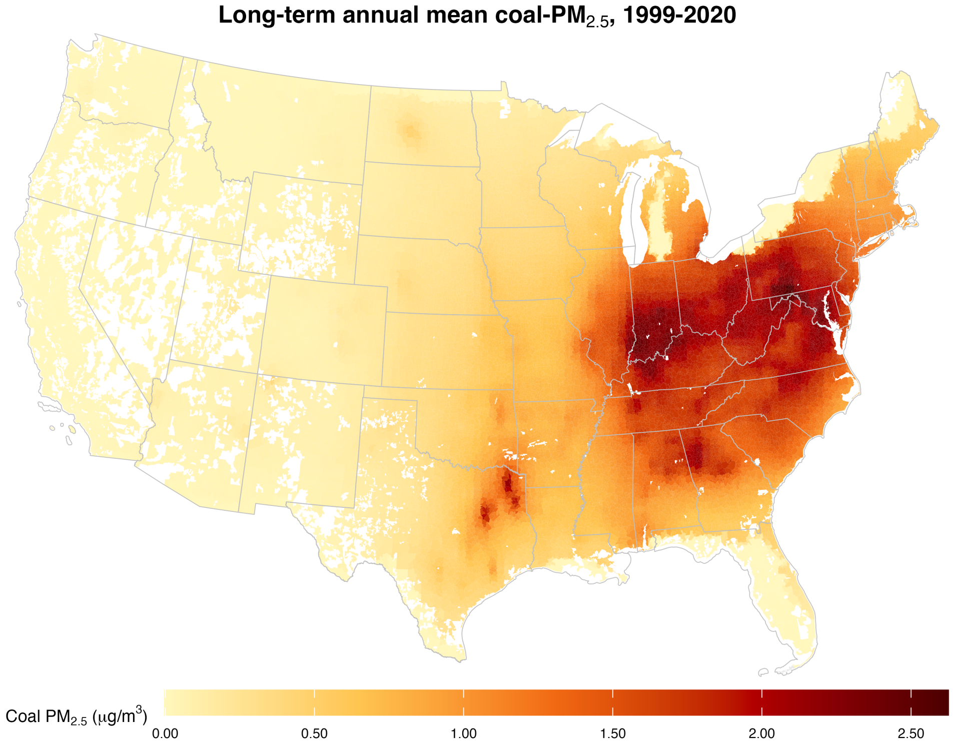


**Fig. S1. Historical coal PM$_{2.5}$ exposure in the United States, 1999–2020.** ZCTA-level long-term annual mean coal-attributable PM$_{2.5}$ concentrations for 1999-2020 aggregated from all coal-fired power plant source contributions.

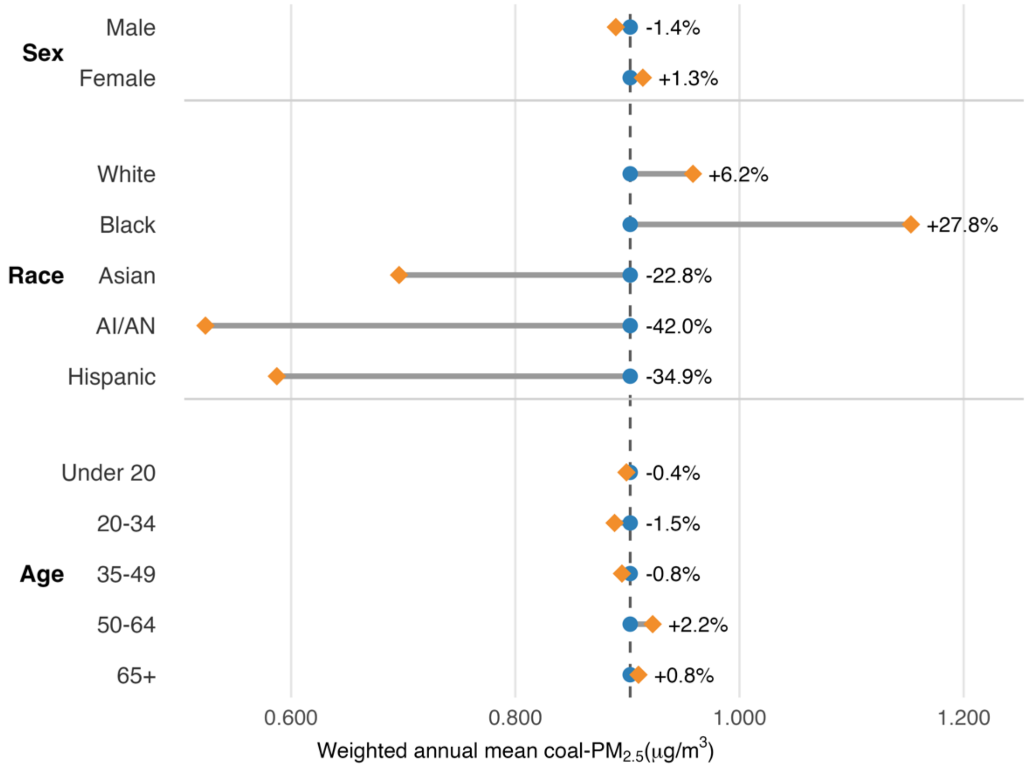


**Fig. S2. Historical population-weighted annual mean coal $PM_{2.5}$ exposure by sex, race and ethnicity, and age group, 1999–2020.** Blue circles indicate the overall population-weighted mean, orange diamonds indicate subgroup-specific population-weighted means, and horizontal segments show the difference between each subgroup mean and the overall mean. Percent labels indicate the percent difference from the overall population-weighted mean.

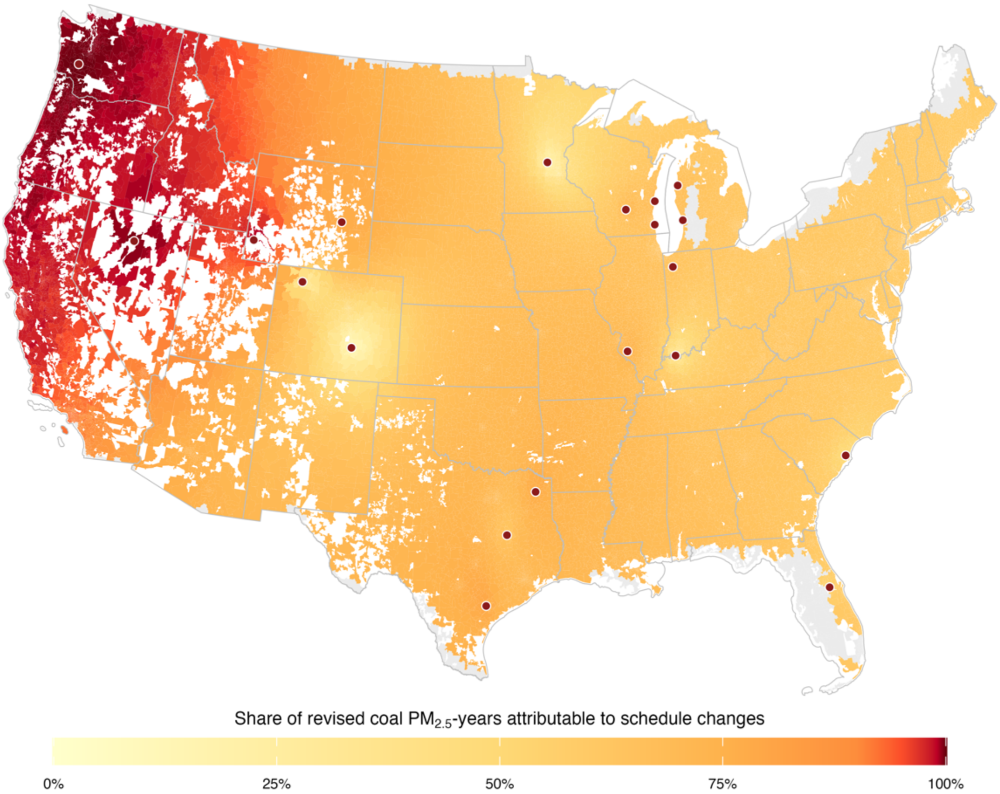


**Fig. S3. Share of cumulative coal $PM_{2.5}$ exposure under the revised retirement scenario attributable to retirement-schedule changes, 2026–2035.** For each ZCTA, the attributable share was calculated as cumulative coal $PM_{2.5}$ exposure under the revised scenario minus cumulative exposure under the original scenario, divided by cumulative exposure under the revised scenario. Values therefore represent the percentage of revised-scenario coal $PM_{2.5}$-years attributable to the retirement-schedule changes among the 20 affected plants. Points indicate plants with later or removed retirement dates. White areas fall outside Census ZCTA coverage; gray ZCTAs had zero or unavailable cumulative exposure under the revised scenario, for which the attributable share was undefined.

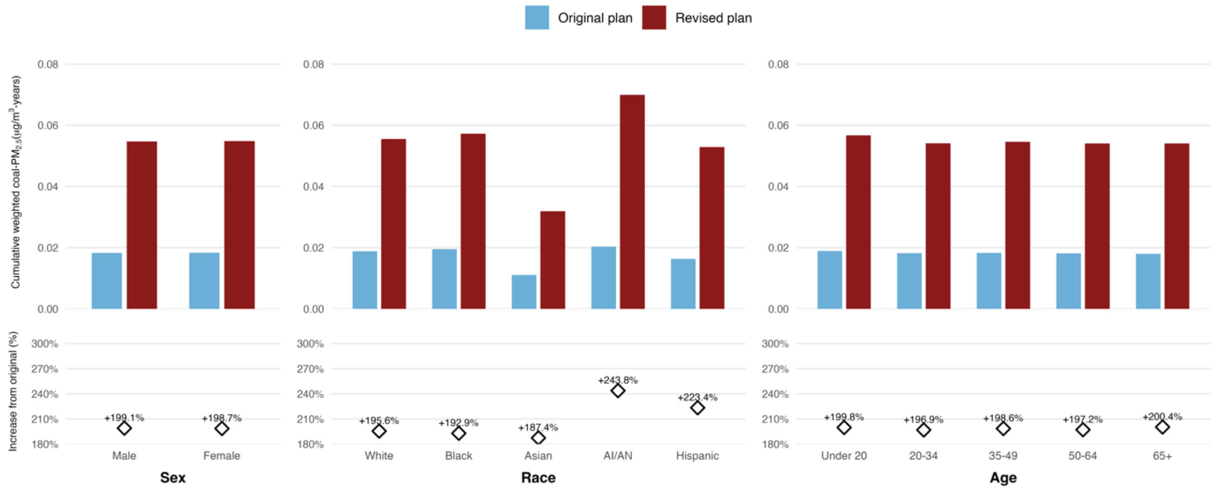


**Fig. S4. Cumulative population-weighted coal $PM_{2.5}$ exposure under original and revised retirement scenarios, 2026–2035.** Cumulative population-weighted coal $PM_{2.5}$ exposure from the 20 affected plants under the original and revised retirement scenarios, by sex, race, and age group. Blue bars show the original retirement scenario and red bars show the revised retirement scenario. Lower panels show the percent increase under the revised scenario relative to the original scenario. Values are reported in micrograms per cubic meter–years. Percent labels indicate the revised-minus-original difference divided by the original scenario.

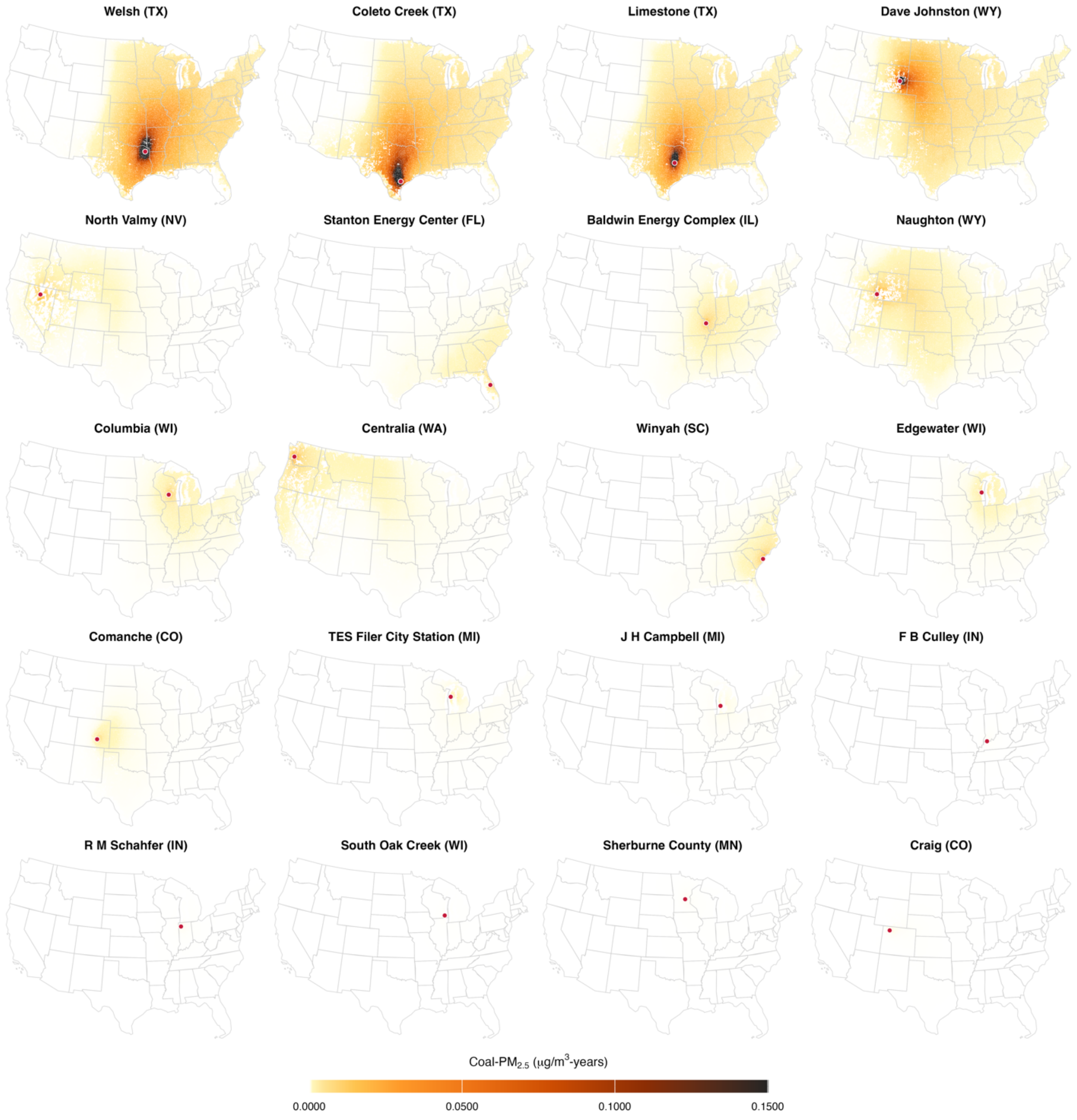


**Fig. S5. Plant-specific cumulative additional coal PM2.5 exposure from delayed or removed retirement dates, 2026–2035.** Maps show ZCTA-level cumulative additional coal PM2.5 exposure under the revised retirement scenario relative to the original retirement scenario for each of the 20 affected plants. Values are summed over 2026–2035 and reported in micrograms per cubic meter–years. Points indicate plant locations.

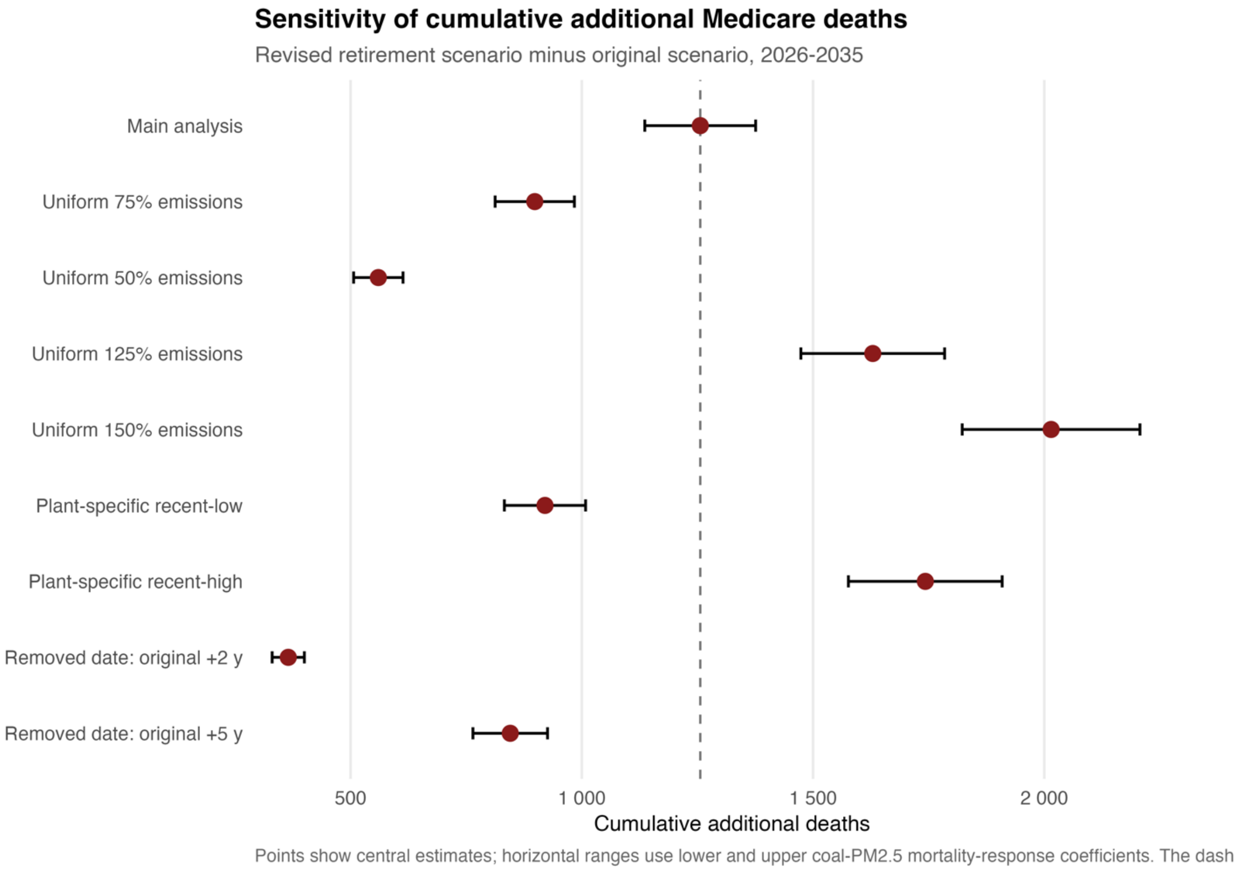


**Fig. S6. Sensitivity of cumulative additional Medicare mortality to alternative future $SO_2$ emissions and retirement-date assumptions, 2026–2035.** Points show central estimates of cumulative additional deaths under the revised retirement scenario relative to the original scenario, and horizontal intervals show lower and upper estimates based on the coal $PM_{2.5}$ mortality-response coefficients. The vertical line indicates the main-analysis estimate of 1,256 additional deaths. Alternative emissions assumptions were applied to both the original and revised retirement scenarios. Alternative retirement dates were assigned at the generator level, only to generators whose retirement date was removed; generators that retained a reported date kept it.

**Table S1. Coal-fired power plants with delayed or removed retirement dates between January 2024 and January 2026 EIA-860M inventories**. Retirement dates are reported per generator in EIA-860M; a plant was classified as having a delayed or removed retirement date if at least one coal-fired generator had a later date, or no reported date, in January 2026. A dash indicates no reported retirement date. Unit-level $SO_2$ is the 2023–2025 mean annual mass reported to EPA CAMPD. "Affected $SO_2$" is the share of the plant's coal $SO_2$ emitted by generators classified as delayed or date removed; plant-level $SO_2$ was prorated by emissions-weighted generator availability.

| Plant | Unit | $SO_2$ (t/yr) | Original retirement date | Revised retirement date | Affected $SO_2$ |
|---|---|---|---|---|---|
| **Limestone (TX)** | 1 | 6,191 | Dec 2029 | — | 100% |
| | 2 | 2,936 | Dec 2029 | — | |
| **Comanche (CO)** | 2 | 964 | Dec 2025 | Dec 2026 | 37% |
| | 3 | 1,623 | — | — | |
| **Stanton (FL)** | 1 | 708 | Dec 2025 | — | 49% |
| | 2 | 737 | — | — | |
| **Baldwin (IL)** | 1 | 1,157 | Dec 2025 | Dec 2027 | 100% |
| | 2 | 1,216 | Dec 2025 | Dec 2027 | |
| **F B Culley (IN)** | 2 | 177 | Dec 2025 | Mar 2026 | 13% |
| | 3 | 1,153 | — | — | |
| **J H Campbell (MI)** | 1 | 2,167 | May 2025 | Feb 2026 | 100% |
| | 2 | 1,430 | May 2025 | Feb 2026 | |
| | 3 | 1,442 | May 2025 | Feb 2026 | |
| **Centralia (WA)** | 2 | 1,039 | Dec 2025 | — | 100% |
| **South Oak Creek (WI)** | 5 | 4 | May 2024 | May 2024 | 94% |
| | 6 | 4 | May 2024 | May 2024 | |
| | 7 | 66 | May 2025 | Dec 2026 | |
| | 8 | 62 | May 2025 | Dec 2026 | |
| **Edgewater (WI)** | 5 | 440 | Jun 2025 | — | 100% |
| **Dave Johnston (WY)** | 1 | 2,031 | Dec 2027 | — | 88% |
| | 2 | 2,270 | Dec 2027 | — | |
| | 3 | 786 | Dec 2027 | Dec 2027 | |
| | 4 | 1,308 | Dec 2027 | — | |
| **Naughton (WY)** | 1 | 566 | Dec 2025 | — | 100% |
| | 2 | 609 | Dec 2025 | — | |
| **Craig (CO)** | 1 | 410 | Dec 2025 | Mar 2026 | 28% |
| | 2 | 312 | Sep 2028 | Sep 2028 | |
| | 3 | 722 | Dec 2029 | Dec 2029 | |
| **R M Schahfer (IN)** | 17 | 242 | Dec 2025 | Jun 2026 | 100% |
| | 18 | 151 | Dec 2025 | Jun 2026 | |
| **Sherburne County (MN)** | 1 | 155 | Dec 2025 | Dec 2026 | 5% |
| | 3 | 2,954 | Dec 2034 | Dec 2034 | |
| **Welsh (TX)** | 1 | 5,762 | Jan 2028 | — | 100% |
| | 3 | 5,627 | Jan 2028 | — | |
| **Coleto Creek (TX)** | 1 | 9,412 | Dec 2027 | — | 100% |
| **Winyah (SC)** | 1 | 449 | Dec 2028 | Dec 2030 | 100% |
| | 2 | 480 | Dec 2028 | Dec 2030 | |
| | 3 | 816 | Dec 2028 | Dec 2030 | |

| | | | | | |
|---|---|---|---|---|---|
| | 4 | 641 | Dec 2028 | Dec 2030 | |
| **Columbia (WI)** | 1 | 810 | Jun 2026 | Dec 2029 | 100% |
| | 2 | 823 | Jun 2026 | Dec 2029 | |
| **North Valmy (NV)** | 1 | 2,290 | Dec 2025 | Dec 2025 | 21% |
| | 2 | 622 | Dec 2025 | — | |
| **TES Filer City Station (MI)** | GEN1 | 128 | Jun 2025 | — | 100% |

**Table S2. Historical comparison of coal plant retirement-schedule revisions across two-year EIA-860M windows**. For each baseline inventory, we defined eligible plants as those with at least one operating coal-fired generator that reported a retirement year earlier than the baseline year plus 11, matching the main analysis criterion of retirement before 2035 for the January 2024 inventory. We used matched plant and generator identifiers in the January inventory two years later to classify later dates and retirement dates removed.

| Comparison period | Eligible plants | Later date | Date removed | Later or removed, n (%) |
|---|---|---|---|---|
| 2016–2018 | 52 | 5 | 6 | 11 (21.2%) |
| 2018–2020 | 33 | 3 | 1 | 4 (12.1%) |
| 2020–2022 | 53 | 5 | 6 | 11 (20.8%) |
| 2022–2024 | 70 | 7 | 5 | 12 (17.1%) |
| 2024–2026 | 50 | 10 | 10 | 20 (40.0%) |

**Table S3. Plant-specific additional Medicare deaths attributable to delayed or removed retirement dates, 2026-2035.** Values are additional deaths under the revised retirement scenario relative to the original retirement scenario, summed across ZCTAs and years. Values in parentheses indicate lower and upper estimates calculated using the lower and upper coal $PM_{2.5}$ mortality-response coefficients.

| Plant | Additional Medicare deaths (lower–upper) |
|---|---|
| Welsh (TX) | 434.7 (393.2–476.2) |
| Coleto Creek (TX) | 316.0 (285.9–346.2) |
| Limestone (TX) | 279.2 (252.6–305.9) |
| Dave Johnston (WY) | 98.7 (89.3–108.1) |
| Baldwin (IL) | 20.1 (18.1–22.0) |
| Naughton (WY) | 17.8 (16.1–19.5) |
| Columbia (WI) | 17.4 (15.8–19.1) |
| Stanton (FL) | 15.9 (14.4–17.4) |
| Centralia (WA) | 14.6 (13.2–16.0) |
| Winyah (SC) | 13.2 (12.0–14.5) |
| Edgewater (WI) | 11.6 (10.4–12.7) |
| North Valmy (NV) | 7.3 (6.6–8.0) |
| TES Filer City Station (MI) | 3.4 (3.1–3.7) |
| Comanche (CO) | 2.5 (2.2–2.7) |
| J H Campbell (MI) | 2.1 (1.9–2.3) |
| R M Schahfer (IN) | 0.6 (0.5–0.6) |
| Sherburne County (MN) | 0.5 (0.4–0.5) |
| South Oak Creek (WI) | 0.4 (0.3–0.4) |
| F B Culley (IN) | 0.3 (0.3–0.3) |
| Craig (CO) | 0.2 (0.2–0.2) |

**Table S4. Model relating $SO_2$ emissions to coal $PM_{2.5}$ concentrations.** The model estimated the historical relationship between plant-level $SO_2$ emissions and ZCTA-level coal $PM_{2.5}$ concentrations using plant-ZCTA-year observations from 1999-2020. The model included plant-ZCTA fixed effects and standard errors clustered by plant. Observations with zero $SO_2$ emissions or zero coal $PM_{2.5}$ were excluded before log transformation. The $SO_2$ elasticity estimate indicates the percent change in coal $PM_{2.5}$ associated with a 1% change in $SO_2$ emissions. Predicted concentrations were multiplied by a Duan smearing factor, mean(exp(residual)), computed separately for each plant.

| | |
|---|---|
| **Model specification** | |
| Outcome | log(coal $PM_{2.5}$) |
| Predictor | log($SO_2$ emissions) |
| Fixed effects | Plant-ZCTA |
| Standard errors | Clustered by plant |
| **Sample** | |
| Years | 1999-2020 |
| Unit of observation | Plant-ZCTA-year |
| Observations | 12,474,346 |
| Plant-ZCTA pairs | 616,801 |
| **Model results** | |
| Gamma | 1.165 |
| Standard error | 0.021 |
| t statistic | 56.287 |
| P value | $<2.2 \times 10^{-16}$ |
| RMSE | 0.538 |
| Adjusted R2 | 0.945 |
| Within R2 | 0.834 |
| **Duan smearing factor** | |
| Duan smearing factor, pooled | 1.169 |
| Duan smearing factor, range across plants | 1.063 – 1.226 |

**Table S5. National Medicare enrollment used to project ZCTA-level Medicare populations, 2010–2035.** Observed enrollment values were obtained from Centers for Medicare & Medicaid Services data, and projected values for 2026–2035 were obtained from the Medicare Trustees Report and accompanying supplementary tables.

| Year | National Medicare enrollment |
|---|---|
| 2010 | 47,663,396 |
| 2011 | 48,892,758 |
| 2012 | 50,768,714 |
| 2013 | 52,425,659 |
| 2014 | 54,013,038 |
| 2015 | 55,496,222 |
| 2016 | 56,981,183 |
| 2017 | 58,457,244 |
| 2018 | 59,989,883 |
| 2019 | 61,514,510 |
| 2020 | 62,840,267 |
| 2021 | 63,892,626 |
| 2022 | 65,100,546 |
| 2023 | 66,509,869 |
| 2024 | 67,601,000 |
| 2025 | 69,537,000 |
| 2026 | 71,101,000 |
| 2027 | 72,778,000 |
| 2028 | 74,402,000 |
| 2029 | 75,888,000 |
| 2030 | 77,134,000 |
| 2031 | 78,105,000 |
| 2032 | 78,945,000 |
| 2033 | 79,711,000 |
| 2034 | 80,520,000 |
| 2035 | 81,349,000 |

**Table S6. National Medicare deaths, enrollment, and baseline all-cause mortality rates, 2010–2023.** Death counts and enrollment values were obtained from Centers for Medicare & Medicaid Services data. Baseline all-cause mortality rates were calculated as deaths among Medicare beneficiaries divided by national Medicare enrollment. ZCTA-level baseline rates for the 2026–2035 projections were calculated from 2019 CMS data.

| Year | Deaths among Medicare beneficiaries | National Medicare enrollment | Baseline all-cause mortality rate |
|---|---|---|---|
| 2010 | 2,003,962 | 47,663,396 | 0.04204 |
| 2011 | 2,046,334 | 48,892,758 | 0.04185 |
| 2012 | 2,077,591 | 50,768,714 | 0.04092 |
| 2013 | 2,126,488 | 52,425,659 | 0.04056 |
| 2014 | 2,144,287 | 54,013,038 | 0.03970 |
| 2015 | 2,220,438 | 55,496,222 | 0.04001 |
| 2016 | 2,226,692 | 56,981,183 | 0.03908 |
| 2017 | 2,292,249 | 58,457,244 | 0.03921 |
| 2018 | 2,323,160 | 59,989,883 | 0.03873 |
| 2019 | 2,337,988 | 61,514,510 | 0.03801 |
| 2020 | 2,750,553 | 62,840,267 | 0.04377 |
| 2021 | 2,750,141 | 63,892,626 | 0.04304 |
| 2022 | 2,656,380 | 65,100,546 | 0.04080 |
| 2023 | 2,517,604 | 66,509,869 | 0.03785 |

**Table S7. Sensitivity of cumulative additional Medicare deaths to alternative future $SO_2$ emissions and retirement-date assumptions, 2026–2035.** Values are additional cumulative deaths among Medicare beneficiaries. Percentage changes are relative to the main analysis. Uniform scaling was applied to every generator; recent-low and recent-high use each generator's minimum and maximum annual $SO_2$ during 2023–2025. Alternative emissions assumptions were applied to both the original and revised retirement scenarios. Alternative retirement dates were assigned only to generators whose retirement date was removed.

| Sensitivity scenario | Additional Medicare deaths | Change |
|---|---|---|
| **Main analysis** | **1,256 (1,136–1,376)** | |
| Uniform 50% emissions | 560 (507–614) | -55% |
| Uniform 75% emissions | 898 (813–984) | -28% |
| Uniform 125% emissions | 1,630 (1,474–1,785) | +30% |
| Uniform 150% emissions | 2,016 (1,823–2,208) | +60% |
| Unit-specific recent-low | 922 (834–1,009) | -27% |
| Unit-specific recent-high | 1,747 (1,580–1,914) | +39% |
| Removed date: original +2 years | 365 (331–400) | -71% |
| Removed date: original +5 years | 846 (765–926) | -33% |

**Table S8. Balancing-authority assignments and hyperscale data-center (HDC) electricity demand for the 20 coal-fired power plants with delayed or removed retirement dates.** Balancing-authority assignments are based on EPA eGRID2023 Revision 2. HDC demand values represent the total annual electricity demand of hyperscale data centers located within each balancing authority under the central facility-load scenario (u=0.58) of Guidi et al. (*36*). A plant was classified as included in the HDC attribution when its balancing authority contained positive HDC demand in the analytical sample.

| Plant | State | EIA plant ID | Balancing authority | Included in HDC attribution? | Central HDC demand in BA (TWh/year) |
|---|---|---|---|---|---|
| **Limestone** | TX | 298 | ERCO | Yes | 4.608 |
| **Comanche** | CO | 470 | WACM | Yes | 1.230 |
| **Stanton Energy Center** | FL | 564 | FMPP | No | 0 |
| **Baldwin Energy Complex** | IL | 889 | MISO | Yes | 7.169 |
| **F B Culley** | IN | 1012 | MISO | Yes | 7.169 |
| **J H Campbell** | MI | 1710 | MISO | Yes | 7.169 |
| **Centralia** | WA | 3845 | BPAT | Yes | 5.406 |
| **South Oak Creek** | WI | 4041 | MISO | Yes | 7.169 |
| **Edgewater** | WI | 4050 | MISO | Yes | 7.169 |
| **Dave Johnston** | WY | 4158 | PACE | Yes | 1.270 |
| **Naughton** | WY | 4162 | PACE | Yes | 1.270 |
| **Craig** | CO | 6021 | WACM | Yes | 1.230 |
| **R M Schahfer** | IN | 6085 | MISO | Yes | 7.169 |
| **Sherburne County** | MN | 6090 | MISO | Yes | 7.169 |
| **Welsh** | TX | 6139 | SWPP | Yes | 6.427 |
| **Coleto Creek** | TX | 6178 | ERCO | Yes | 4.608 |
| **Winyah** | SC | 6249 | SC | No | 0 |
| **Columbia** | WI | 8023 | MISO | Yes | 7.169 |
| **North Valmy** | NV | 8224 | NEVP | Yes | 1.651 |
| **TES Filer City Station** | MI | 50835 | MISO | Yes | 7.169 |
